# The effect of smaller classes on infection-related school absence: Evidence from the Project STAR randomized controlled trial

**DOI:** 10.1101/2021.05.16.21257297

**Authors:** Paul T. von Hippel

## Abstract

In an effort to reduce viral transmission, many schools are planning to reduce class size if they have not reduced it already. Yet the effect of class size on transmission is unknown. To determine whether smaller classes reduce school absence, especially when community disease prevalence is high, we merge data from the Project STAR randomized class size trial with influenza and pneumonia data from the 122 Cities Mortality Reporting System on deaths from pneumonia and influenza.

Project STAR was a block-randomized trial that followed 10,816 Tennessee schoolchildren from kindergarten in 1985-86 through third grade in 1988-89. Children were assigned at random to small classes (13 to 17 students), regular-sized classes (22 to 26 students), and regular-sized class with a teacher’s aide.

Mixed effects regression showed that small classes reduced absence, but not necessarily by reducing infection. In particular, small classes reduced absence by 0.43 days/year (95% CI -0.06 to -0.80, *p*<0.05), but had no significant interaction with pneumonia and influenza mortality (95% CI -0.27 to +0.30, *p*>0.90). Small classes, by themselves, may not suffice to reduce the spread of viruses.

## INTRODUCTION

In response to the pandemic of COVID-19 (coronavirus disease 2019), caused by the novel virus SARS-CoV-2 (severe acute respiratory syndrome coronavirus 2), many US schools implemented policies to reduce class size. During the 2020-21 school year, approximately one-half of schools implemented some type of “hybrid” instruction policy that reduced in-person class size by having a substantial fraction of children stay home to receive instruction remotely.^1^ Although more than three-quarters of US adults are projected to be vaccinated when schools reopen in fall 2021,^2^ there is still concern about fully reopening schools at class sizes that were typical before the pandemic. Out of the $125 billion allocated to public^3^ and private^4^ elementary and secondary schools under the American Rescue Plan passed in March 2021, the Biden administration, advised by the American Federation of Teachers, has suggested spending $50 billion to hire new teachers and reduce class sizes by approximately 10 percent.^5^

The rationale for class size reduction is straightforward. With fewer children in a classroom, there is a lower probability that the classroom will contain an infected child, and fewer children to whom an infected child will be exposed. Children can sit or stand further apart, at least on average, reducing the chance of infection by direct contact or through the air. With fewer children to supervise, teachers and staff can devote more time and attention to disinfecting surfaces and enforcing healthy behaviors, such as wearing masks and washing hands. A simulation published in the pandemic summer of 2020 predicted that doubling class size could more than double the size of COVID-19 outbreaks.^6,7^

Empirically, though, the evidence regarding the effect of class size on infection is limited. In the US, two observational studies have estimated the effect of school reopening policies during the pandemic—fully online, fully in-person, or hybrid—on the incidence of COVID-19 hospitalizations^8^ and positive COVID-19 tests^9^ in fall 2020. Although hybrid reopening implies smaller average class sizes than full reopening, both studies found that hybrid reopening had no significant effect in counties where the prevalence of COVID-19 before schools reopened was low. In counties where the prior prevalence of COVID-19 was high, the results were mixed; one study concluded that fully opening schools in-person accelerated the spread of the virus,^9^ while the other reported ambiguous results that were sensitive to model specification.^8^ During fall 2020 many counties had high prevalence, but by fall 2021 most counties are expected to have low prevalence and a population that is largely vaccinated, especially in the most vulnerable groups. Under these circumstances, the results suggest that schools can fully reopen, with safeguards, with little risk of re-sparking the pandemic.

Evidence on the effect of class size on infection is limited not just for the novel coronavirus (SARS-CoV-2), but also for more familiar pathogens such as influenza. While correlations between class size and infection are sometimes reported in observational studies,^10^ we have found only one working paper that tried to estimate the causal effect of reducing class size on influenza-related absence.^11^ Exploiting discontinuities induced by Japanese laws limiting class size, the study concluded that reducing Tokyo class sizes to 27 from an average of 32 would have substantially reduced the risk of school closures due to outbreaks during the flu seasons of 2015-2017.^11^ Below a class size of 27, the benefits of further reductions were less clear. Note that most US classes are already smaller than 27 students; average US class size is 17 to 26, depending on grade level and class type.^12^

In this study, we estimate the effect on influenza-related absence of reducing average class size from 23 to 15. We use evidence from a randomized controlled trial that assigned young US children to larger and smaller classes at random. The trial started many years before the advent of SARS-CoV-2, but against the usual backdrop of seasonal pneumonia and influenza. This naturally reduces the study’s relevance to the current COVID-19 pandemic, but it also gives the study several strengths.

One strength is that class size, along with presence of a teacher’s aide, was the only variable manipulated in the trial. This eliminates the problem of separating the effect of class size from the effects of other school and community policies. Another strength is that class size was manipulated at random—something that schools have not done during the COVID-19 pandemic—so that the effect of class size can be interpreted as unambiguously causal.

An additional strength is that, schools and school-age children may play only a small role in the COVID-19 pandemic,^13,14^ school-age children clearly play a major role in both transmitting and manifesting symptoms of influenza. Indeed, school-age children “drive” influenza prevalence, playing a larger role than any other age group in transmitting influenza viruses.^15^ During influenza pandemics, incidence tends to spike after schools open^16^ and subsides, at least among school-age children, when schools close for two weeks or more.^17,18^ In addition, while children rarely display symptoms of SARS-CoV-2, school-age children often have influenza symptoms so severe that they stay home from school.^19^ If reducing class size has a substantial effect on influenza transmission, therefore, the effect should be evident in reduced rates of absence from school, at least in communities where the prevalence of influenza is high.

## METHODS

### Project STAR randomized controlled trial

Our primary data come from Tennessee’s Student/Teacher Achievement Ratio Project (Project STAR)—a four-year block-randomized longitudinal trial funded by a $12 million appropriation from Tennessee’s House Bill 544, passed by the Tennessee State Legislature in May 1985.^20–23^ Data from Project STAR are anonymized and publicly available, and thus exempt from institutional review for human subjects research.

Within each school that participated in Project STAR, children and teachers were assigned at random to three classroom treatments in kindergarten: (1) small classes with a target size of 13 to 17 students, (2) regular-sized classes with a target size of 22 to 26 students, or (3) regular-sized class with a teacher’s aide. Children were followed from kindergarten in 1985-86 through third grade in 1988-89. Children who spent kindergarten in a small class remained in small classes from kindergarten through third grade. Children who spent kindergarten in one of the other conditions—a regular-sized class or a regular-sized class with an aide—were re-randomized between those two conditions after kindergarten. Children who entered participating schools after kindergarten were randomized among the three conditions as well.

180 of the 886 elementary schools in Tennessee volunteered for Project STAR.^20^ 79 schools were selected to participate in kindergarten, and a total of 80 schools participated over the four years of the study, with little attrition. Schools were selected with the goal of achieving geographic diversity and the requirement that participating schools have at least 57 kindergartners, enough to populate at least one classroom in each of the three experimental conditions—*i*.*e*., at least one classroom with 13-16 students and at least two classrooms with 22 to 26 students each (13+22+22=57).

We use Project STAR to estimate the effect of smaller classes and teachers’ aides on absence. Number of days absent from school was recorded for participating children for three of the four study years: kindergarten 1985-86, first grade 1986-87, and third grade 1988-89, but not second grade 1987-88. As far as we know, ours is the first study to use Project STAR to estimate the effect of small classes on absence. The original purpose of Project STAR was to estimate the effects of reducing class size on test scores in grades K-3,^20–23^ and later analyses also estimated effects on grade repetition, high school graduation, college attendance and completion, and early adult employment and wages.^24,25^

### 122 Cities Mortality Reporting System (CMRS)

Although Project STAR recorded the number of days absent each year, it did not record how many absences were due to illness. This is a common limitation. Even today, school data rarely specify the reason for absence; at most, systems distinguish between absences that are excused or unexcused. Because of data limitations, there are few studies of reasons for absence, but the studies that exist suggest that approximately half of school absences are due to illness.^26–28^ Past efforts to estimate infection-related absence sometimes relied on correlations between absence among schoolchildren and disease prevalence in the larger community.^29^ That is the strategy that we adopt here.

To estimate the correlation between absence and infection, we merged Project STAR with data from the 122 Cities Mortality Reporting System (CMRS), a surveillance study run by the Centers for Disease Control and Prevention from 1962 through 2016.^30^ The CMRS recorded mortality data for 122 US cities, including the four largest cities in Tennessee: Memphis, Nashville, Knoxville, and Chattanooga. For some cities, the data include the surrounding metropolitan area; for others, they are limited to the city proper. The CMRS did not cover other areas of Tennessee that participated in Project STAR.

For each city and week, the CMRS recorded the total number of deaths, as well as the number of deaths that were due to pneumonia and influenza (PI). Deaths were reported for each week, and we aggregated them to each school year. The school year was defined as running from week 34 of one year to week 22 of the next. For example, deaths during the kindergarten school year of 1985-86 were defined as the total of deaths from week 34 of 1985 (starting August 24) through week 22 of 1986 (starting May 31). Changing the beginning and end of the school year by a few weeks would not materially change the results, since the vast majority of PI deaths were concentrated in December and January.

For each city and school year, we calculated PI mortality—the percentage of deaths that were due to PI. PI mortality is often interpreted as a proxy for the prevalence and virulence of influenza viruses.^31^ Although school-age children rarely die of PI, children miss school more often during weeks when PI mortality peaks among the elderly and other vulnerable adults.^29^ Although we would have liked to have separate estimates of PI mortality for the attendance zone of each school, PI mortality was only available at the city level. This is a common limitation in infectious disease surveillance, which is often aggregated to larger geographic areas such as cities, counties, or multi-county regions.^8,9^

### Data merging

To merge the CMRS with the Project STAR data, we had to identify the location of each Project STAR school. The Project STAR data do not identify schools explicitly, but we identified them by merging with other data. In particular, Table II in the Project STAR *Technical Report*^32^ listed the name and district of all 80 participating schools, along with the number of small classes, regular classes, and regular classes with an aide that each school offered in each year of Project STAR. Other characteristics of the Project STAR were available in the US Department of Education’s Common Core of Data,^33^ which provided data on every US school back to 1986-87 (year 2 of Project STAR). By matching the Project STAR data to variables from the Technical Report and Common Core, we identified which schools in the Project STAR were in the four cities surveyed by the CMRS, and this gave us the community PI mortality for those schools. Because of differences between data sources, matches between the CMRS and Project STAR were approximate but not exact. We used the *ultimatch* command for Stata to minimize the Euclidean distance between matched schools.^34^ Alternative matching procedures yielded practically identical estimates; in the few cases where the matched school differed, the matched district was the same, and so was the community PI mortality.

### Statistical analysis

Because Project STAR assigned children and teachers to treatments at random, we could have estimated the effect of class size simply by comparing the average number of absences in each treatment group. However, getting efficient point estimates with accurate standard errors required careful model specification. Because the effect of class size was small, we maximized power by pooling data longitudinally across the years of the study. In analyzing the pooled data, we had to account for correlations among observations of the same child in different years, as well as correlations among different children in the same classroom and school year.

To estimate the average effect of class size on absence, we fit the following mixed model in Stata software:

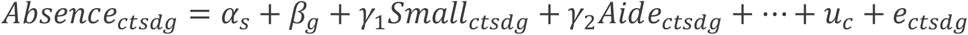

Here *Absence*_ctsdg_ was the number of absences for child *c* with teacher *t* in school *s* and district *d* during grade *g* (kindergarten 1985-86, first grade 1986-87, or third grade 1988-89). *small*_*ctsdg*_ and *Aide*_*ctsdg*_ indicated which experimental treatment the child received during that grade—a small class or a regular class with an aide; regular classes without an aide were the reference. *α*_*s*_ and *β*_*g*_ were school and grade fixed effects, which account for time trends and the fact that children were randomized to conditions within schools. *u*_*c*_ is a child random effect used to model the correlation among observations of the same child in different grades. e_ctsdg_ is a random residual, clustered at the classroom level to account for the correlation among observations of different children in the same classroom. We estimated the model using the *xtreg* command in Stata software, version 16.1.

Because of random assignment, the experimental treatments were not correlated with any child characteristics, so no child-level covariates were needed to get unbiased estimates of treatment effects. Nevertheless, we fit the model both with and without covariates representing each child’s race, gender, and free lunch eligibility (an indicator of poverty). Unsurprisingly, these covariates changed the results very little.

To estimate whether the effect of class size on absence was stronger in communities and years with higher infection rates, we added a covariate *PI*_*sg*_, representing PI mortality in district *d* during the school year when the child was in grade *g*. We centered *PI*_*sg*_ around its mean of 7.3, and we let the mean-centered variable interact with the experimental treatments:

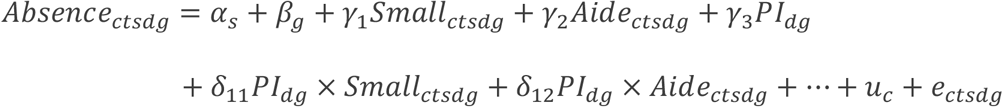

Although community infection rates could not be randomized, the coefficients of *PI*_*dg*_ and the interactions can be interpreted as causal effects if the year and school fixed effects control adequately for unobserved confounding variables that vary between schools and years. Again, we fit the model both with and without covariates for race, gender, and free lunch eligibility. Again, the covariates made little difference to the results.

## RESULTS

Table 1 summarizes the design of Project STAR and the characteristics of participating children, schools, and school districts. The table is limited to children with absence data in kindergarten, first, or third grade, since no absence data was recorded in second grade. In each of those three school years, over 6,000 children participated in over 300 classrooms; across all three years, over 10,000 distinct children participated in nearly 1,000 classrooms. Approximately equal numbers of classrooms were assigned to the three experimental treatments, but because the small classrooms had fewer students, slightly less than one-third of children were assigned to the small class condition. Nearly two-thirds of participating children were white, one-third were black, and less than one percent were other races and ethnicities. Just under half of participating children were female, and precisely half were poor enough to receive free school lunches. Comparisons elsewhere show that Project STAR students were poorer and more likely to be black than children in in other Tennessee schools and other states in the 1980s.^24^

**Table 1.** Description of Project STAR randomized controlled trial, Tennessee, school years 1985-89.

|  | 1985-86 | 1986-87 | 1988-89 | All 3 years |
| --- | --- | --- | --- | --- |
| <u>Outcome</u> |  |  |  |  |
| Absences / year (mean) | 10.5 | 7.6 | 6.8 | 8.3 |
| <u>Treatments (randomly assigned)</u> |  |  |  |  |
| % small class | 30 | 28 | 32 | 30 |
| % regular-sized class | 35 | 38 | 31 | 34 |
| % regular-sized class with teacher's aide | 35 | 33 | 38 | 35 |
| <u>Covariates</u> |  |  |  |  |
| % female | 49 | 48 | 48 | 48 |
| % free lunch | 48 | 52 | 50 | 50 |
| % white | 67 | 66 | 66 | 66 |
| % black | 32 | 33 | 34 | 33 |
| % other race/ethnicity (incl. Hispanic, Asian, Native American) | 0.5 | 0.8 | 0.5 | 0.6 |
| <u>Sample size</u> |  |  |  |  |
| Observations | 6,251 | 6,662 | 6,586 | 19,499 |
| Distinct students | 6,251 | 6,662 | 6,586 | 10,816 |
| Teachers (classrooms) | 325 | 337 | 331 | 993 |
| Schools | 79 | 76 | 74 | 80 |
| <u>Number of schools by district</u> |  |  |  |  |
| Memphis Public Schools | 20 | 19 | 19 | 20 |
| Knox County Public Schools (incl. Knoxville) | 5 | 4 | 4 | 5 |
| Davidson County Public Schools (incl. Nashville) | 4 | 4 | 4 | 4 |
| Hamilton County Public Schools (incl. Chattanooga) | 3 | 3 | 3 | 3 |
| Other school districts | 47 | 46 | 44 | 48 |

The bottom of Table 1 shows the distribution of participating schools across Tennessee school districts. About one-third of participating schools were in the four cities covered by the CMRS. Fully a quarter of participating schools were in Memphis, while another 15 percent were in Knox County (principally Knoxville), Davidson County (principally Nashville), and Hamilton County (principally Chattanooga).

The top of Table 1 shows the average number of absences per year, which dropped from over 10 in kindergarten 1985-86 to less than 7 by third grade 1988-89. Figure 1 compares absences across the three experimental treatments. The differences were small but consistent across kindergarten, first, and third grade, with each year having fewer absences in small classes than in regular-sized classes or regular-sized classes with a teachers’ aide. Table 2 shows that these differences are statistically significant (*p*<.05), with smaller classes having 0.4 fewer annual absences per student, on average, across kindergarten, first, and third grade. Including gender, race, and free lunch status as covariates had practically no effect on this result.

**Table 2.** Mixed model predicting days of absence per year, Project STAR, Tennessee, school years 1985-89.

|  | Coef. | 95% CI | Coef. | 95% CI | Coef. | 95% CI | Coef. | 95% CI |
| --- | --- | --- | --- | --- | --- | --- | --- | --- |
| Small class | -0.43* | -0.80, -0.06 | -0.42* | -0.79, -0.05 | -0.49+ | -1.01, 0.04 | -0.43 | -0.95, 0.09 |
| Teacher's aide | 0.21 | -0.13, 0.54 | 0.20 | -0.14, 0.54 | 0.20 | -0.28, 0.68 | 0.19 | -0.29, 0.67 |
| First grade (ref. kindergarten) | -2.70** | -3.00, -2.41 | -2.73** | -3.03, -2.43 | -1.94** | -2.42, -1.46 | -1.97** | -2.44, -1.50 |
| Third grade (ref. kindergarten) | -3.33** | -3.64, -3.03 | -3.36** | -3.66, -3.06 | -3.38** | -4.39, -2.37 | -3.46** | -4.43, -2.48 |
| PI |  |  |  |  | 0.38* | 0.00, 0.75 | 0.39* | 0.02, 0.76 |
| Small class × PI |  |  |  |  | 0.02 | -0.27, 0.30 | 0.02 | -0.27, 0.30 |
| Teacher's aide × PI |  |  |  |  | -0.00 | -0.28, 0.27 | -0.01 | -0.28, 0.26 |
| Female (ref. male) |  |  | 0.26+ | -0.03, 0.55 |  |  | 0.23 | -0.20, 0.65 |
| Black (ref. white) |  |  | -1.48** | -2.06, -0.90 |  |  | -1.19** | -2.06, -0.33 |
| Other race/ethnicity |  |  | -1.65* | -3.30, -0.00 |  |  | -2.10* | -3.83, -0.37 |
| Free lunch |  |  | 1.64** | 1.34, 1.93 |  |  | 1.86** | 1.40, 2.32 |
| Observations | 19,499 |  | 19,329 |  |  | 8,073 |  | 8,028 |
| Distinct children | 10,816 |  | 10,726 |  |  | 4,966 |  | 4,936 |

*Note.* \*\* $p < 0.01$ , \* $p < 0.05$ , + $p < 0.1$ , two sided. Coef.=Regression coefficients. CI=classroom-clustered 95 percent confidence intervals. PI=percent of deaths due to pneumonia and influenza, mean-centered. All models include school fixed effects, year fixed effects, and child random effects.

**Figure 1.**
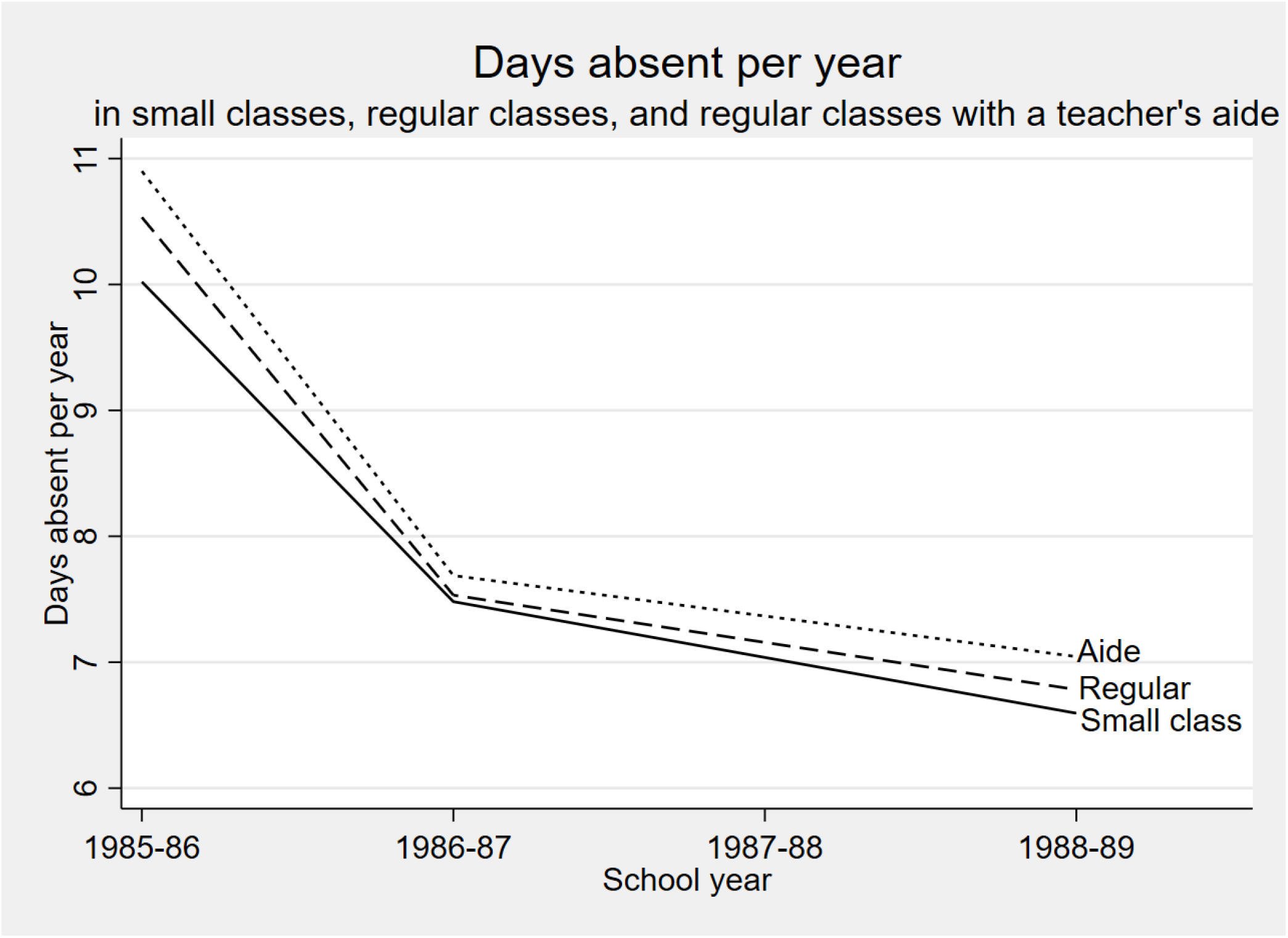
Smaller classes had slightly fewer days absent, on average, than regular-sized classes or regular-sized classes with a teacher’s aide.

Although these results show that smaller classes reduced absence, it is not clear whether the reduction in absence was due to a reduction in infection. To address that question, we added community PI mortality to the model for the cities covered by the CMRS. Figure 2 summarizes trends in PI across the four cities. In Nashville, PI mortality held steady between 5 and 6 percent across the 4 years of Project STAR. In Memphis, PI mortality rose from 6 to nearly 10 percent, and in Knoxville and Chattanooga, PI mortality rose from approximately 7 to approximately 9 percent. The differences in levels and trends within and between cities help to identify the effect of PI mortality on absence.

**Figure 2.**
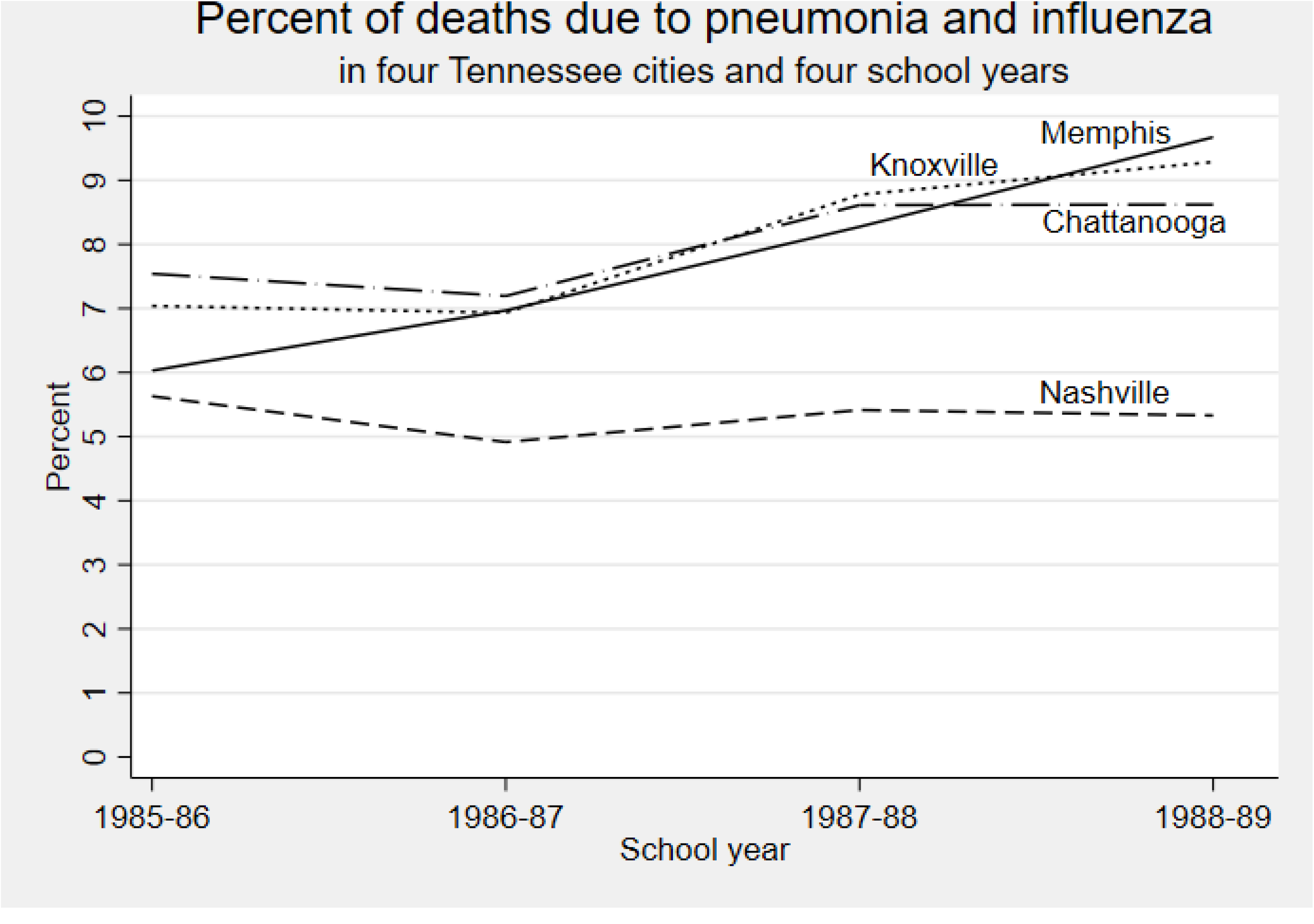
Percent of deaths due to pneumonia and influenza in four Tennessee cities and four school years.

Table 2 shows that PI mortality was a significant predictor (*p*<.05) of absence; a one percentage point increase in PI mortality was associated with an increase of approximately 0.4 annual absences per student. However, community PI rates did not appear to moderate the effect of class size on absence. The interaction between PI and the small class size condition was near zero and far from statistical significance (*p*>0.9). The interaction between PI and the teacher’s aide condition was also near zero and far from statistical significance (*p*>0.9).

Including PI and interactions in the model slightly reduces the statistical significance of the effect of small classes. The point estimate for the small class effect is similar in all models, but the confidence interval gets slightly wider and the p values get slightly larger when PI and interactions are included in the model (from *p*=0.02 to *p*=0.06 without covariates, from *p*=0.03 to *p*=0.11 with covariates). This is partly because the sample size is reduced to districts with PI data and partly because there is some correlation between the small class variable and its interaction with PI.

## DISCUSSION

In Tennessee’s Project STAR randomized controlled trial, reducing class sizes by one-third reduced annual absence by 0.4 days per child on average. Variation in absence rates were correlated with variation in PI mortality across cities and years, but the effect of class size on absence was no greater when PI mortality was high than it was when PI mortality was low. Although smaller classes reduced absence, it is not clear that they did so by reducing infection. They might have done so by other mechanisms—such as increasing teachers’ engagement with children and parents—but the data offer no measures that can be used to test other mechanisms.

The analogy to the COVID-19 pandemic is of course imperfect. Influenza and COVID-19 are different viruses, and Project STAR did not take place during a pandemic. During the COVID-19 pandemic, class size reduction was usually part of a multi-pronged strategy that included measures that were not used during Project STAR, such as mask-wearing, regular disinfection of surfaces, and avoidance of mass assemblies during recess and lunch. As the COVID-19 pandemic recedes, though, the analogy will improve, as mask mandates are dropped and school assemblies resume, while federal funds continue to be available for class size reduction through September 2023.

Nevertheless, our results do suggest some lessons relevant to the COVID-19 pandemic. School-age children play a much larger role in transmitting influenza than in transmitting COVID-19, so the fact that class-size reduction did little to reduce PI-correlated absences in Project STAR suggests that it might do even less to slow the spread of COVID-19. Project STAR reduced class sizes by one-third, which is less than the 50 percent reductions that some hybrid schools enacted during the height of the pandemic, but far more than the 10 percent reduction that the White House called for in its February 2021 proposal.^5^ Our results raise the possibility that a 10 percent reduction in class size may not be effective in reducing the prevalence of COVID-19. And if vaccination is effective and widespread, class size reduction may not be necessary as a public health measure.

## Data Availability

Data and code needed to replicate the results are available on the website of the Open Science Foundation.

https://osf.io/scg6e/

## ACKNOWLEDGMENTS

The author(s) did not seek funding for this research.

